# Inflammatory dietary potential is associated with vitamin depletion and gut microbial dysbiosis in early pregnancy

**DOI:** 10.1101/2023.12.02.23299325

**Authors:** Suzanne A. Alvernaz, Elizabeth S. Wenzel, Unnathi Nagelli, Lacey B. Pezley, Bazil LaBomascus, Jack A. Gilbert, Pauline M. Maki, Lisa Tussing-Humphreys, Beatriz Peñalver Bernabé

## Abstract

**Background:** Pregnancy alters many physiological systems, including the maternal gut microbiota. Diet is a key regulator of this system and can alter the host immune system to promote inflammation. Multiple perinatal disorders have been associated with inflammation, maternal metabolic alterations, and gut microbial dysbiosis, including gestational diabetes mellitus, preeclampsia, preterm birth, and mood disorders. However, the effects of high inflammatory diets on the gut microbiota during pregnancy have yet to be fully explored.

**Objective:** To use a systems-based approach to characterize associations among dietary inflammatory potential, a measure of diet quality, and the gut microbiome during pregnancy.

**Methods:** Forty-nine pregnant persons were recruited prior to 16 weeks of gestation. Participants completed a food frequency questionnaire (FFQ) and provided fecal samples. Dietary inflammatory potential was assessed using the Dietary Inflammatory Index (DII) from FFQ data. Fecal samples were analyzed using 16S rRNA amplicon sequencing. Differential taxon abundance with respect to DII score were identified, and microbial metabolic potential was predicted using PICRUSt2.

**Results:** Inflammatory diets were associated with decreased vitamin and mineral intake and dysbiotic gut microbiota structure and predicted metabolism. Gut microbial compositional differences revealed a decrease in short chain fatty acid producers such as *Faecalibacterium,* and an increase in predicted vitamin B12 synthesis, methylglyoxal detoxification, galactose metabolism and multi drug efflux systems in pregnant individuals with increased DII scores.

**Conclusions:** Dietary inflammatory potential was associated with a reduction in the consumption of vitamins & minerals and predicted gut microbiota metabolic dysregulation.

## Introduction

In pregnancy, an under or over supply of nutrients can have deleterious impacts on both maternal and fetal health. For instance, lack of adequate folic acid intake during pregnancy is one of the leading causes of neural tube defects during fetal development (1). Similarly, iron utilization increases during the course of pregnancy, and inadequate supply is associated with poor fetal outcomes, including intrauterine growth restriction and low birth weight (2). Conversely, oversupply of dietary nutrients, including carbohydrates and saturated fats, common in Western diets, are associated with chronic inflammation and can lead to obstetric complications, from gestational diabetes mellitus (GDM) (3,4) to preterm birth (5). This is especially important for minoritized women of color who may have poor nutritional intake due to structural inequalities (6,7,8) and consequently, higher burden of adverse pregnancy outcomes (9). Thus, understanding the pro-inflammatory nature of diets could serve to reduce negative obstetrics and delivery outcomes (10).

Diet is a major regulator of the gut microbiota (11,12). The gut microbiota encompasses the bacteria, fungi, viruses, and protists living inside the human gastrointestinal tract. It is estimated that the combined genomes of all gut bacteria comprise >5 million genes (13), with the potential to metabolize a vast number of different substrates. Over or under supply of dietary nutrients (such as fats or fiber) can provide competitive advantages or disadvantages for different gut microbial species based on their individual metabolic capabilities (14,15). The dynamic nature of pregnancy alters almost every system in the body, including the maternal gut microbiota (16) and immune system (17), which adapts in a tightly regulated clock to maintain immune protection of the mother while simultaneously avoiding autoimmune rejection of the growing fetus (17,18). The structure of the gut microbiota changes as the pregnancy progresses (19–21). In fact, transplantation of gut microbiota from pregnant individuals into germ-free animals, renders common pregnancy phenotypes of obesity, insulin resistance (19,22), and adaptations in immunity (23). Poor diet quality leading to a pro-inflammatory state can alter the normal dynamic changes of the gut microbiota (14) and immune system during pregnancy (24), increasing risk of common perinatal complications, including GDM (25), iron deficiency (26), and mood disorders (27). It is thus essential to understand how maternal diet quality during pregnancy impacts the gut microbiota.

The Dietary Inflammatory Index (DII) is a literature-derived population-based index to quantify the inflammatory potential of diets among diverse populations (28). The Index was developed by leveraging global dietary studies to assign inflammatory effect scores (*S)* to common dietary nutrients based on their ability to increase or decrease pro-inflammatory biomarkers, such as cytokines IL-1β, IL-4, IL-6, and IL-10 (28–30). Previous studies have shown DII is positively associated with inflammatory markers during pregnancy (31), increased rates of cesarean delivery in obese mothers (32) and decreased fetal growth (33). Furthermore, DII has also been negatively linked with microorganisms that produce short-chain fatty acids (SCFAs), which are beneficial anti-inflammatory metabolites (34–37). Thus, the normal gut microbial compositional changes occurring during the gestational period may be negatively altered by poor diet quality, which could be assessed by the DII score, and may mediate obstetric complications.

Dysbiosis refers to the imbalance in the gut microbiota, where the equilibrium between beneficial and harmful microorganisms is disrupted by factors such as poor diet, antibiotics, or illness (25). Microbial dysregulation, closely related to dysbiosis, describes when the regulation of these microbial populations is disturbed, leading to health issues (27) which can manifest as an impaired immune response or altered metabolic processes (21). Both dysbiosis and microbial dysregulation are crucial concepts in understanding conditions like obesity and gestational diabetes, where the gut microbiota plays a significant role in disease progression or mitigation (21). Understanding how diet regulates the gut microbiota during pregnancy could potentially lead to avenues of early interventions to reduce risk of pregnancy comorbidities associated with systemic inflammation. Here, we aim to assess the relationship between dietary inflammatory potential and the maternal gut microbiota during the first trimester of pregnancy in a cohort mostly composed of minoritized women of color living in a large diverse urban community in the United States.

## Methods

### Participant Recruitment

This work is a secondary data analysis of a longitudinal cohort study (MoMent) in which participants were recruited from the outpatient obstetrics clinics at a public university hospital, the University of Illinois Chicago (Chicago, IL, USA), from 2018 to 2020 (38). This study was approved by the University of Illinois Chicago Institutional Review Board (IRB #2014-0325). Written informed consent was obtained prior to study enrollment and sample collection. To be eligible for the study, participants had to be less than 16 weeks pregnant and English speaking. Women were excluded for the following criteria: less than 18 or over 64 years of age, current multi-gestational pregnancy, a prior history of gastrointestinal surgeries, oral antibiotic, antiviral, or antifungal use in the last 6 months, use of medication or supplements to treat any chronic disorder (e.g., diabetes, hypertension, mood disorders), history of substance abuse (excluding marijuana, alcohol and tobacco, self-report) within the last 6 months, use of *in vitro* fertilization treatments for current pregnancy, active diagnosis of cancer, HIV or eating disorders or chronic diarrhea within the last 6 months. For this secondary study, we selected participants who completed a diet food frequency questionnaire before 28 gestational weeks and provided a fecal sample at their first study visit (< 16 gestational weeks), rendering a total of 49 subjects.

### Stool Collection

Study participants self-collected rectal swabs (n=44), avoiding touching the rectal tissue, or provided stool samples (n=5) for gut microbiota assessment. Stool samples were homogenized and aliquoted in cryogenic vials. Rectal swabs and aliquoted stool samples were stored at −80°C before being sent for 16S rRNA amplicon sequencing. Biological samples were collected with an average estimated gestational age of 10.9 ± 3 weeks.

### Dietary Assessment

Participants completed one of two validated FFQs: Vioscreen (n=25) (39) or the Diet History Questionnaire II (DHQII) (n=24) (40) with an average estimated gestational age of 14.7 ± 5.9 weeks. Participants were asked about the previous month of intake. Vioscreen was completed electronically at home by participants, with some receiving calls from research staff to complete the survey. DHQII was completed in-person with a certified registered dietitian within an average of 4.4 ± 5.5 weeks of microbiome sample collection. The Dietary Inflammatory Index (DII) was calculated using the DII components common to both FFQs, a total of twenty-seven variables (60% of total DII parameters) which is within the DII’s developer’s suggested limit (28). Individuals were checked to ensure daily caloric intake < 500 or > 5,000 kcal/day). These DII variables included were daily intake of alcohol (g), vitamin B12 (μg), vitamin B6 (mg), β-carotene (μg), caffeine (g), carbohydrates (g), cholesterol (mg), energy (kcal), total fat (g), fiber (g), folic acid (μg), iron (mg), magnesium (mg), monounsaturated fatty acids (myristoleic acid, MUFA 14:1) (g), niacin (mg), total protein (g), polyunsaturated fatty acids (PUFA) (g), riboflavin (mg), saturated fat (g), selenium (μg), thiamin (mg), trans-saturated fat (g), vitamin A (retinol equivalents), vitamin C (mg), vitamin D (μg), vitamin E (mg), and zinc (mg). Individual DII scores were calculated using (eq. 1):

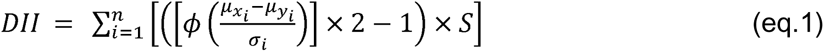

where *n* represents the total number of common DII parameters between VioScreen and DHQII; *µ*_xi_ is the mean daily intake of food parameter *i* obtained from the FFQ; *µ*_yi_ is the global mean (average daily intake across global populations) and; *σ*_i_ is the global standard deviation of parameter *i* both derived from the reference table; *ϕ* is the cumulative distribution function; and *S* represents the inflammatory effect score. Scores can range from −8.87 to +7.98 with the latter being the most inflammatory (28). After calculating DII scores for each participant, individuals were grouped into tertiles. Differences in patient demographics by DII tertile were assessed using Chi-square (qualitative) or ANOVA (quantitative). Correlations among DII parameters (continuous scale) were identified using Spearman’s correlation using energy corrected nutritional values. Energy correction was performed by scaling each individual’s food parameter by their reported daily caloric intake. Dimensionality reduction Principal Component Analysis (PCA) was performed on DII parameters to identify the key nutrients that drive DII scores. Differences in nutrient parameters by tertile were assessed using ANOVA and between Tertile 1 & Tertile 2/3 using students t-test. All analysis were completed in R.

### Microbiota Assessment

Rectal and fecal samples underwent 16S rRNA amplicon sequencing in four different batches at the University of Chicago (Chicago, IL, US) and at the University of California San Diego (San Diego, CA, US) together with control samples to account for possible reactant and environmental contaminations. Forward raw FASTQ sequences were processed using the DADA2 pipeline independently using default parameters (41) and passed to the R package *phyloseq* (42). After primer removal, reads were truncated to 150 base pairs, denoised using standard parameters, and chimeras were removed. Taxonomical assessment of the trimmed, cleaned reads was performed using the Silva reference database version 132 (43). Contaminating amplicon sequence variances (ASV) found in blank controls were removed from each batch using the prevalence method in the R *decontam* package (44). A threshold of 0.5 was used to identify contaminants that were more prevalent in negative controls than in clinical samples. Samples with library size below 10 reads were excluded from downstream analysis. Subsequently, batch-effects were removed using the R package *ComBat-seq* (45). The count table and taxonomic assignments for each batch were then merged, keeping all the Amplicon Sequencing Variants (ASVs). ASVs with a relative abundance less than 1% relative to sample library size were removed from downstream analysis. After prevalence filtering, taxa counts were normalized using cumulative sum scaling (CSS) (46). Alpha diversity was calculated using the Shannon (47) and Simpson indexes (48). Statically significant differences in mean alpha diversity between DII tertiles were assessed using Wilcoxon rank sum test (49). Beta diversity was determined with Bray-Curtis (50) and unweighted, normalized UniFrac distance (51). Significant differences in beta-diversity distances by DII scores were assessed using PERMANOVA (52) correcting for participant BMI, gestational weeks (EGA), food frequency questionnaire type (DHQII or Vioscreen), sample type (stool or rectal) and maternal age. Associations between DII and CSS-normalized ASVs were identified by fitting a zero-inflated Gaussian model with the R package *metagenomeSeq* (53). Models were adjusted by the same covariates as before. Multiple comparisons were corrected using the Benjamini-Hochberg method (54). Finally, gut metabolic potential was predicted via PICRUSt 2.0 (Phylogenetic Investigation of Communities by Reconstruction of Unobserved States) (55). Associations between metabolic pathways, microbial enzymes and DII scores were assessed with zero-inflated Gaussian models, corrected by the same covariates as above and multiple comparisons were adjusted using the Benjamin-Hochberg’s method. Gene set enrichment analysis (GSEA) was performed using all microbial enzymes, identified as significant before FDR adjustment using the R package *MicrobiomeProfiler* (56). Finally, associations among the identified enzymes and each food parameter used in DII estimation were quantified with zero-inflated Gaussian models, corrected by the same covariates as above and multiple comparisons were adjusted using Benjamin-Hochberg’s method. A total of 27 models were fit with Z-scored energy corrected food parameters per subject as the outcome and microbial enzymes as predictors.

## Results

### Our sample was composed of minoritized women of color with a large percentage consuming a vitamin depleted pro-inflammatory diet

A total of 49 participants completed a FFQ and provided a fecal sample. The study cohort was primarily comprised of non-Hispanic Black (44%) and Hispanic (17%) pregnant persons with an average estimated gestational age of 10.9 ± 3 weeks at fecal sample collection, average maternal age of 29 ± 6 years, and 73% reporting an annual household income below $31,000 per year (**Table 1**). Notably, most participants reported use of Federal Aid Health Insurance (75.5%), a proxy for low socioeconomic status (57). A similar number of participants completed the Vioscreen (n=25) and DHQII (n=24) FFQs. Based on the 27 food parameters common between both FFQs (60% of total DII parameters), DII scores were spread across low and higher inflammatory scores with the lowest tertile (Tertile 1) mean of −2.3 (± 0.9) and highest (Tertile 3) mean DII of 3.4 (± 0.5) (**Table 1**). All DII scores were within the normal limits specified by the DII score authors (29). Socio-demographic characteristics were similar across all three groups (**Table 1**, p > 0.05). A less inflammatory diet was associated with higher vitamin B12, B6, A, niacin, iron, and zinc. (**Table 2**, p < 0.05). These DII parameters were positively associated with each other (**Figure 1**, p < 0.05). Of the nutrients used to calculate the DII score, the biggest contributors were those negatively associated with DII (**Figure 1**, p < 0.05).

**Figure 1:**
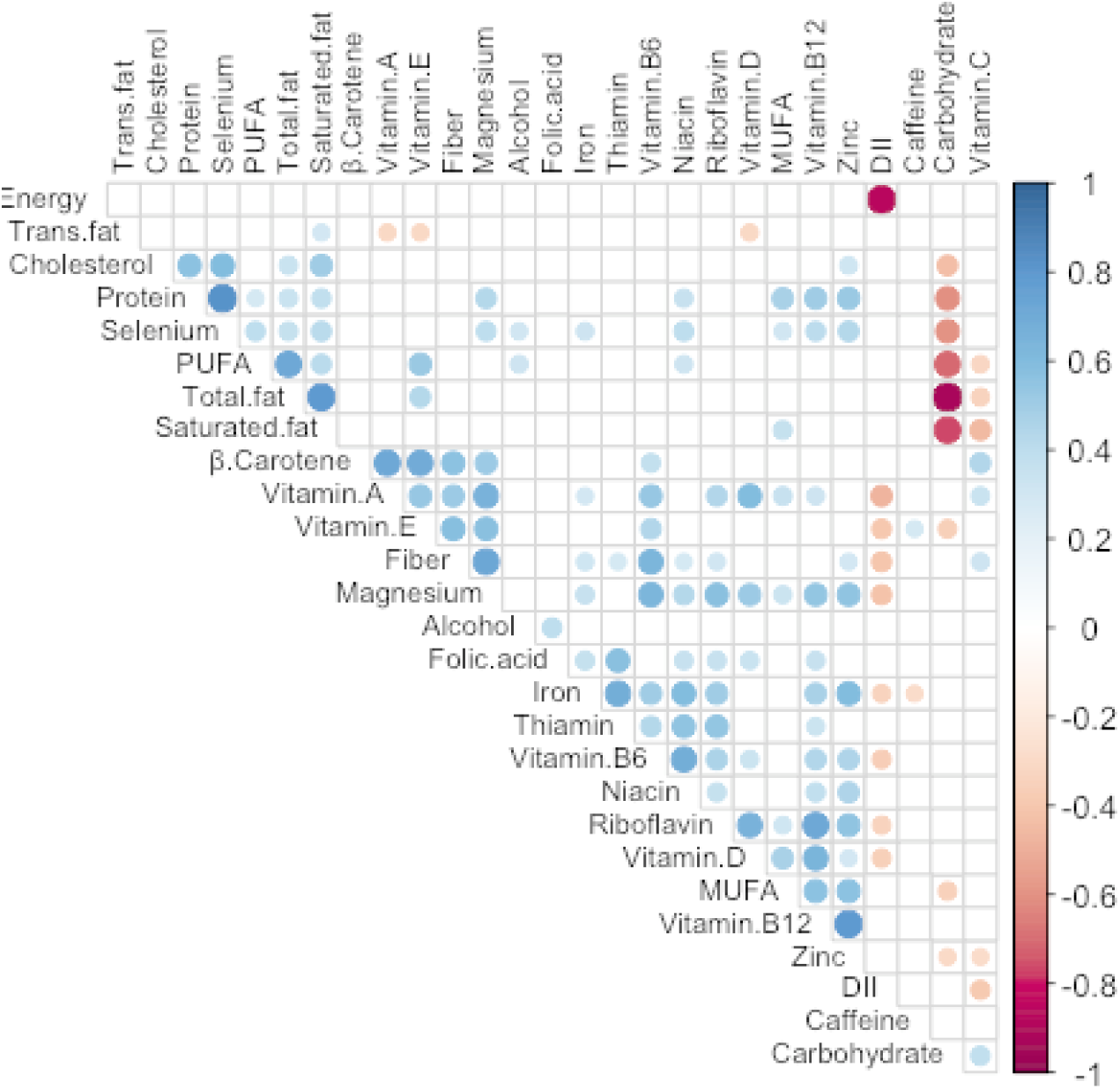
Components and drivers of DII scores. Spearman correlation (p < 0.05) among the 27 parameters used to calculate the DII scores for each subject. Dot size is proportional to the absolute correlation coefficient. See supplemental methods for more details (units and references). PUFA: polyunsaturated fatty acids; MUFA: monounsaturated fatty acids.

**Table 1:** Study cohort demographic characteristics did not differ as a function of DII scores. Participants were stratified into DII tertiles. There were no differences in study characteristics by DII tertile (p > 0.05).

|  |  | **Tertile 1** | **Tertile 2** | **Tertile 3** | **p** |
| --- | --- | --- | --- | --- | --- |
| DII | Mean (SD) | -2.3 (0.9) | 1.0 (0.9) | 3.4 (0.5) |  |
| Gestational Weeks | Mean (SD) | 11.2 (3.5) | 11.2 (2.9) | 10.2 (2.7) | 0.59 |
| Age | Mean (SD) | 28.7 (7.8) | 28.6 (5.3) | 30.2 (5.3) | 0.69 |
| BMI | Mean (SD) | 29.0 (7.2) | 30.4 (6.6) | 28.3 (7.5) | 0.7 |
| Race/Ethnicity | Hispanic | 2 (12.5) | 4 (25.0) | 2 (11.8) | 0.35 |
|  | Non-Hispanic Black | 10 (62.5) | 6 (37.5) | 6 (35.3) |  |
|  | Other/Unreported | 4 (25.0) | 6 (37.5) | 9 (52.9) |  |
| Health Insurance | Federal Aide | 14 (87.5) | 10 (62.5) | 13 (76.5) | 0.26 |
|  | Private | 2 (12.5) | 6 (37.5) | 4 (23.5) |  |
| Education | Above College | 1 (6.2) | 4 (25.0) | 5 (29.4) | 0.34 |
|  | Below College | 6 (37.5) | 4 (25.0) | 7 (41.2) |  |
|  | College | 9 (56.2) | 8 (50.0) | 5 (29.4) |  |
| Employment | Employed Part/Full Time | 11 (68.8) | 8 (50.0) | 10 (58.8) | 0.56 |
|  | Unemployed | 5 (31.2) | 8 (50.0) | 7 (41.2) |  |
| Income | \$31-76k | 2 (12.5) | 3 (18.8) | 2 (11.8) | 0.78 |
| | \$76k+ | 1 (6.2) | 3 (18.8) | 2 (11.8) | |
| | <\$31k | 13 (81.2) | 10 (62.5) | 13 (76.5) | |
| Relationship Status | Married/Relationship | 7 (43.8) | 11 (68.8) | 11 (64.7) | 0.3 |
|  | Single | 9 (56.2) | 5 (31.2) | 6 (35.3) |  |
| Planned Pregnancy | No | 6 (37.5) | 1 (6.2) | 2 (11.8) | 0.17 |
|  | Unreported | 8 (50.0) | 10 (62.5) | 11 (64.7) |  |
|  | Yes | 2 (12.5) | 5 (31.2) | 4 (23.5) |  |

**Table 2:** Differences in nutritional intake by DII tertile. Reported mean (SD) nutrient values were normalized by total Energy intake per day (kcal/day). Vitamin A was reported in retinol equivalents (RE).

|  | **Tertile 1** | **Tertile 2** | **Tertile 3** | **p** |
| --- | --- | --- | --- | --- |
| Alcohol (g) | 0.1 (0.3) | 0.1 (0.2) | 0.1 (0.5) | 0.83 |
| Vitamin B12 (µg) | 3.2 (1.1) | 2.9 (1.2) | 2.2 (0.9) | **0.03** |
| Vitamin B6 (mg) | 1.1 (0.4) | 1.0 (0.3) | 0.8 (0.2) | **0.02** |
| β Carotene (µg) | 1983.5 (1563.7) | 1547.3 (1698.9) | 1491.2 (1104.8) | 0.58 |
| Caffeine (g) | 0.0 (0.0) | 0.0 (0.0) | 0.0 (0.0) | 0.43 |
| Carbohydrate (g) | 127.5 (25.0) | 129.0 (28.0) | 129.1 (33.9) | 0.98 |
| Cholesterol (mg) | 136.6 (66.6) | 145.8 (64.5) | 163.6 (113.8) | 0.65 |
| Energy (kcal) | 2988.0 (924.1) | 1801.1 (409.1) | 969.8 (333.8) |  |
| Total fat (g) | 39.5 (9.6) | 40.3 (9.5) | 39.9 (11.4) | 0.98 |
| Fiber (g) | 10.9 (2.4) | 10.1 (3.3) | 8.9 (2.2) | 0.11 |
| Folic acid (µg) | 168.4 (84.9) | 165.8 (86.0) | 128.7 (75.9) | 0.31 |
| Iron (mg) | 9.1 (3.4) | 7.9 (2.8) | 6.2 (1.7) | **0.01** |
| Magnesium (mg) | 171.2 (35.5) | 147.1 (35.7) | 143.2 (40.8) | 0.08 |
| MUFA (g) | 0.0 (0.0) | 0.1 (0.0) | 0.0 (0.0) | 0.49 |
| Niacin (mg) | 11.7 (3.1) | 10.1 (2.1) | 9.0 (2.8) | **0.02** |
| Protein (g) | 39.4 (6.1) | 35.6 (7.0) | 35.6 (11.4) | 0.37 |
| PUFA (g) | 8.2 (2.3) | 7.9 (2.7) | 8.1 (4.7) | 0.97 |
| Riboflavin (mg) | 1.3 (0.4) | 1.1 (0.4) | 0.9 (0.3) | 0.02 |
| Saturated fat (g) | 13.5 (3.5) | 14.2 (4.1) | 13.4 (4.3) | 0.82 |
| Selenium (µg) | 52.2 (10.2) | 49.5 (11.6) | 51.1 (19.5) | 0.87 |
| Thiamin (mg) | 0.9 (0.3) | 0.8 (0.2) | 0.7 (0.2) | 0.11 |
| Trans fat (g) | 1.7 (0.5) | 1.8 (0.6) | 1.6 (0.7) | 0.75 |
| Vitamin A (RE) | 523.2 (192.7) | 437.0 (191.1) | 358.8 (150.4) | **0.04** |
| Vitamin C (mg) | 85.5 (42.4) | 71.7 (47.2) | 64.5 (53.1) | 0.45 |
| Vitamin D (µg) | 3.9 (1.9) | 3.6 (2.0) | 2.6 (1.7) | 0.1 |
| Vitamin E (mg) | 5.1 (2.1) | 3.9 (1.2) | 4.1 (1.8) | 0.13 |
| Zinc (mg) | 6.5 (1.8) | 5.9 (1.4) | 5.1 (1.4) | **0.04** |

### Gut microbiota composition and predicted metabolic potential were associated with proinflammatory diets in early pregnancy

There were no statistically significant differences in alpha or beta diversity by DII tertile (**Fig. S1A-D**, p-value > 0.05). A total of 18 ASVs were identified as differentially abundant in terms of DII score (**Table S1,** false discovery rate (fdr)-adjusted p-value < 0.05). Among the top 10 ASVs, those mapped to *Solobacterium moorei*, *Gemella asaccharolytica, Gardnerella vaginalis, Atopobium vaginae* and unclassified members of the Eggerthellaceae family and the *Corynebacterium* genera, were positively associated with DII (**Fig. 2A**, adjusted p < 0.05), while those mapped to *Parabacteroides distasonis,* unclassified members of the genus *Faecalibacterium, Prevotella,* and *Clostridium sensu stricto* (**Fig. 2A**, adjusted p < 0.05) were negatively associated with dietary inflammatory potential.

**Figure 2:**
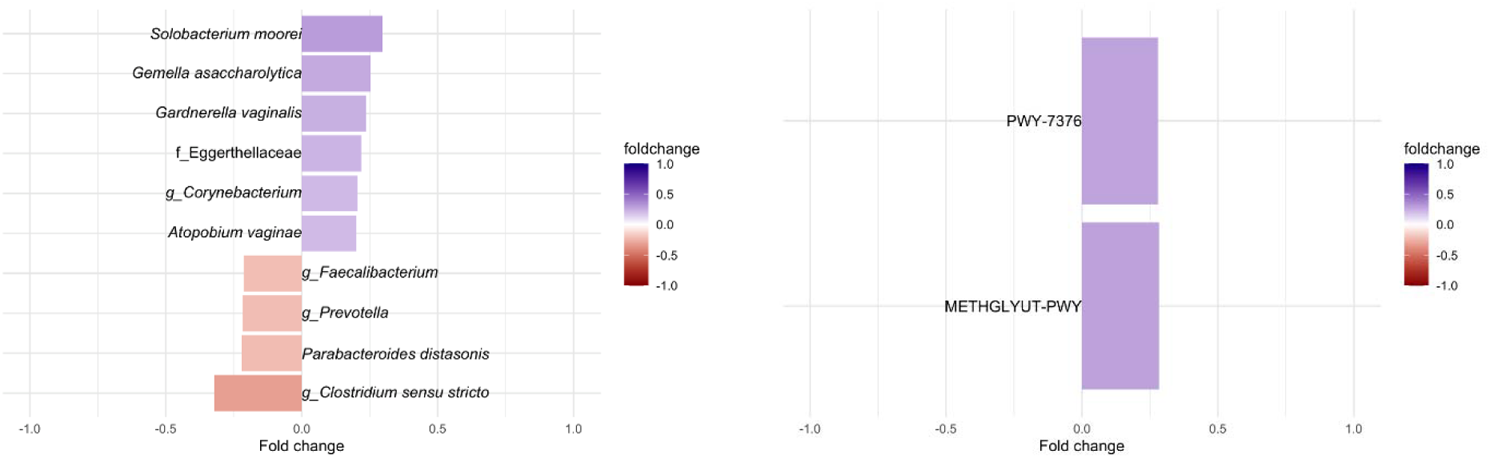
Differentially abundant gut taxa and predicted gut produced enzymes as a function of DII scores. Top 10 CSS normalized taxa (A) and all predicted pathways (B) that were identified as statistically significant differentially abundant by DII after correction by participant age, estimated gestational weeks (EGA), BMI, and food frequency questionnaire type (DHQII or VioScreen), and sample type (adjusted p < 0.05 & adjusted p < 0.1). Taxa names are lowest identifiable rank. Full list of enriched ASVs can be found in **Table S1**. *PWY-7376*: Cob(II)yrinate a,c-diamide biosynthesis II; *METHGLYUT-PWY*: methylglyoxal detoxification super pathway.

Next, we examined which PICRUSt2 predicted microbial enzymes and metabolic pathways were associated with DII scores. We identified 2 pathways, aerobic adenosylcobalamin (vitamin B12) synthesis and methylglyoxal detoxification (**Fig. 2B**, fdr-adjusted p < 0.05), and 38 enzymes significantly associated with DII score (**Table S2,** fdr-adjusted p-value < 0.05). The significantly enriched predicted enzymes were all positively associated with DII **(Table S2**) with several being involved in bacterial two-component system related to multi-drug efflux pumps (*K07642,* BaeS) and drug efflux pumps/resistance (*K18889, K18148*) and in galactose degradation and transport (*K10111, K12112, K0894*) (**Fig. 3A**, fdr-adjusted p < 0.05). Gene set enrichment analysis of the DII associated predicted enzymes before multiple comparisons (n=194, p-value<0.05), also revealed an increase of two-component systems terms (58) primarily related to nitrogen and sugar metabolism, genes involved in nitrogen metabolism (specifically nitrate reduction to ammonia), biofilm formation, and galactose metabolism (**Fig. 3B**, adjusted p < 0.05, **Table S3**).

**Figure 3:**
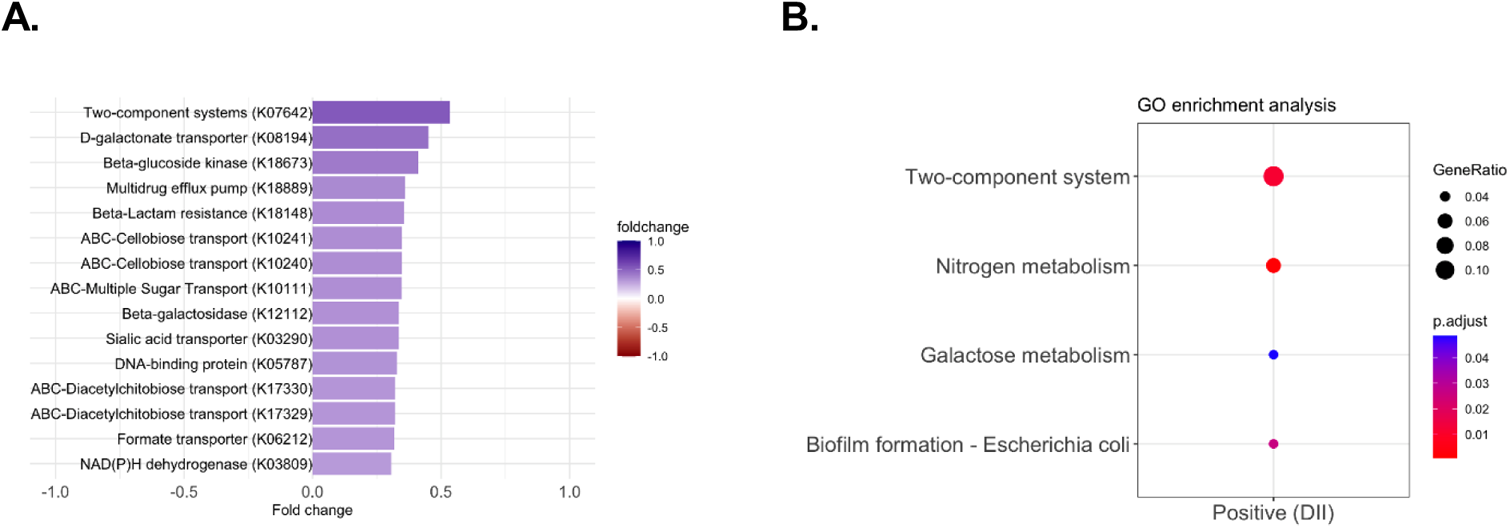
Predicted microbial gene sets enrichment analysis in terms of DII scores. A: Top 15 predicted enzymes that were identified as differentially abundant by DII (adjusted p < 0.05). B: Gene set enrichment of enzymes grouped by those positively (N=194, p < 0.05) associated with DII score. Full list of enriched enzymes can be found in **Table S2**. Full list of enzymes by gene set term can be found in **Table S3**.

### Several individual DII components were associated with predicted microbial enzymes

Finally, we investigated the relationships between DII components and DII-associated enzymes (**Fig. 4**). Several DII-associated enzymes, such as efflux pumps and resistance genes, and enzymes pertaining to the galactose metabolism, were also associated with 19 individual DII food parameters including Vitamins B12, A, D, E, and cholesterol. The microbial resistance genes were *K18889* (multi-drug efflux pump), *K18148* (beta lactamase resistance) and *K07642* (two-component signaling system for efflux pumps). *K07642* was associated with the largest number of DII components (63%, such as vitamins A, C, D, E among others). These enzymes were mostly negatively associated with essential vitamins and minerals (vitamin A, B12, Niacin, & Zinc) that were decreased in the higher DII individuals (**Table 2**, **Fig. 4**). The second group of enzymes associated with individual DII nutrient parameters (Sugar transporter *K10111,* Beta-galactosidase *K12112,* Beta-glucoside kinase *K18673,* and D-galactonate transporter *K01894)* were involved in galactose metabolism and were mostly negatively associated with key perinatal nutrients such as magnesium and folic acid. Cholesterol was the only nutrient positively associated with more than one enzyme (D-galactonate transporter *K01894* & Two-component systems *K07642*)

**Figure 4:**
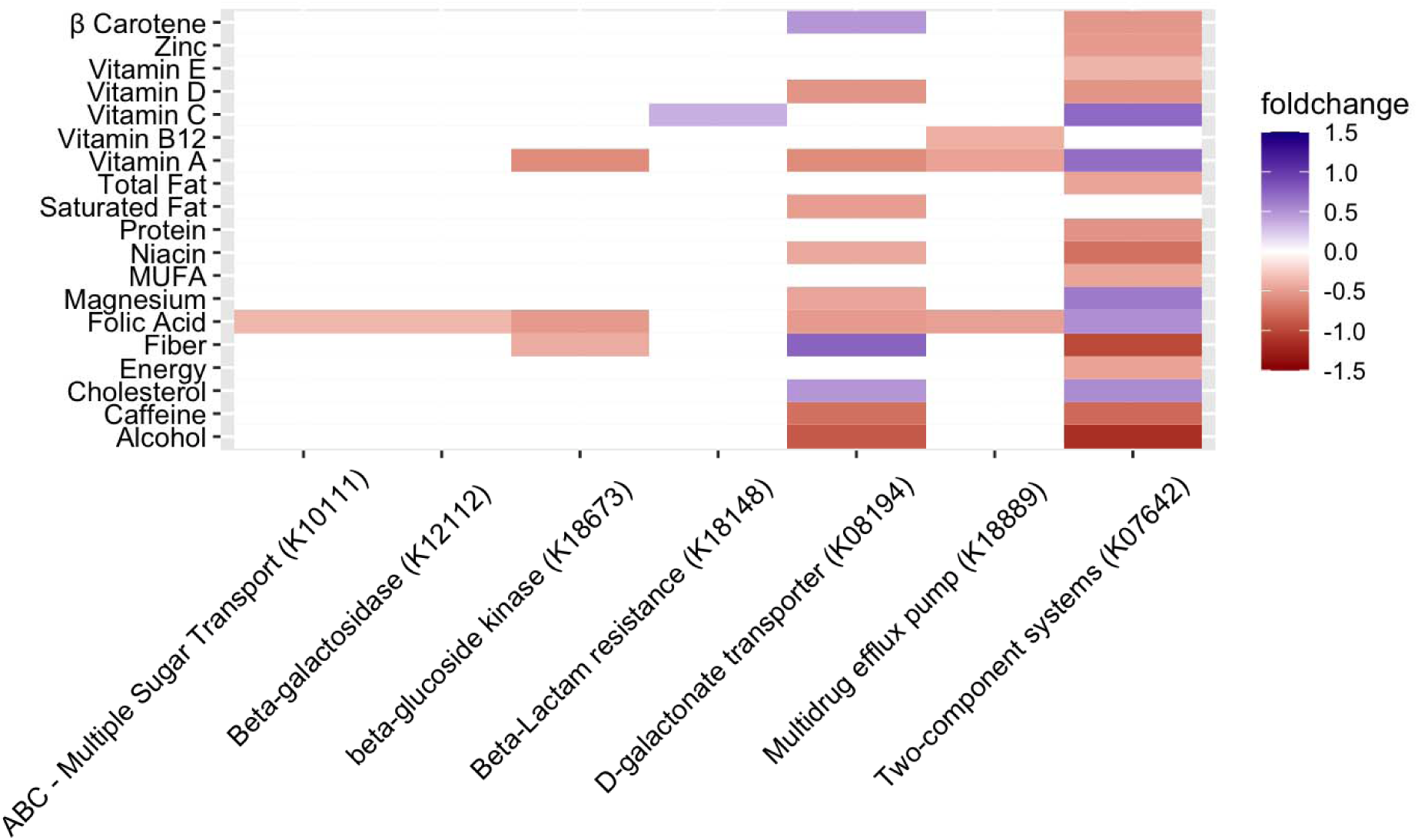
Relationship between predicted enriched enzymes and dietary components. Differential abundance of top 15 predicted enzymes and the 27 dietary components of the DII (adjusted p < 0.05). Only significant associations are represented.

## Discussion

Dietary intake is an essential aspect of maternal health. Food choice is often related to the dietary preferences of an individual, their environment, and their socioeconomic status. Under or oversupply of certain nutrients can have direct impacts on maternal health and the growing fetus (59, 60). This study demonstrated that diet inflammatory potential, an indicator of poor diet quality, was associated with lower vitamin and mineral intake, altered maternal gut microbiota composition and dysregulated microbial metabolic potential in early pregnancy. As diet is one of the main regulators of the gut microbiota (11, 12), poor diet quality during pregnancy could disrupt the normal dynamic adaptations of the maternal gut microbiota through altered substrate availability.

In our study, the overall gut microbiota diversity did not differ in individuals consuming higher inflammatory diets. While distinct patterns of beta diversity composition in pregnant individuals with better diet quality have been previously reported (61, 62), recent microbiome-pregnancy cohorts have not identified alterations in beta diversity by diet quality (63,64), supporting our study observations. At the taxonomic level, several ASVs varied with dietary inflammatory potential. Higher DII scores were associated with enrichment of pro-inflammatory bacterial species, including *S. moorei,* a producer of proinflammatory sulfur compounds (65), and those associated with inflammatory perinatal conditions such as preterm birth and GDM including *G. vaginalis, A. vaginae* (66) and members of the *Corynebacterium* genera (67). In contrast, microbiome members that were depleted in individuals reporting high DII scores included known producers of anti-inflammatory SCFAs such as *Faecalibacterium* (68). This suggests pro-inflammatory diets are associated with deleterious alterations to gut microbiota composition.

The influence of maternal diet quality on the gut microbiota extends to their metabolic potential, as our study reveals a link between the predicted metabolic capabilities of gut microbes in individuals with higher inflammatory diets and community-wide metabolic dysregulation. The Cob(II)yrinate a,c-diamide biosynthesis metabolic pathway (part of adenosylcobalamin/vitamin B12 pathway) (69) was increased in participants reporting higher DII scores. Vitamin B12 deficiency can lead to upregulation of the cytokine TNF-α (70) and has been linked to multiple perinatal disorders including pre-eclampsia and neonate growth retardation (71). The increase in this bacterial pathway may be related to the insufficient vitamin B12 intake of the high DII group and a subsequent shift towards microbial communities capable of producing this essential vitamin to compensate for the unbalance. The second pathway associated with high DII scores was a microbially regulated methylglyoxal detoxification pathway. Methylglyoxal is a toxic oxidizing substance derived from sugar metabolism, a DII enriched process in this study, and is known to be elevated in perinatal metabolic disorders such as gestational diabetes mellitus (72,73). Methylglyoxal detoxification can occur via glyoxalase system (74,75), a common microbial detoxification pathway (76,77). This finding highlights the pro-inflammatory nature of poor diet quality as well as the compensatory shift in the gut microbiota to reduce toxic metabolic species.

DII scores were also associated with the upregulation of microbial virulence pathways, such as drug resistance, biofilm formation as well as nitrogen and sugar/galactose metabolism. Sugar and galactose metabolism overall was enriched in individuals reporting high DII scores. Galactose metabolism has been shown to be enriched in perinatal inflammatory conditions such as gestational diabetes (78,79) and specifically associated with elevated methylglyoxal (80). Notably, *S. moorei* and *G. vaginalis* were both positively linked with DII scores and have been reported to contribute to galactose fermentation (65,81). Enrichment of microbial multidrug resistant efflux pumps enzymes (*K07642,18889, K18148*) could be promoted by host pro-inflammatory diets. Recent work has shown bacterial multidrug efflux pumps are involved in nutrient signal processing, cellular adaptations to anaerobic respiration, and colonization of eukaryotic cells (82). Poor maternal diet quality may promote expression of these gut microbial enzymes in response to nutrient alterations. The predicted gut microbial enzymes related to both galactose metabolism and virulent efflux pumps were also mostly negatively correlated with vitamins and minerals (i.e., vitamins B12 and A and iron, magnesium, niacin, zinc) that were decreased in high DII individuals. Taken together, our results suggest that a vitamin and mineral depleted perinatal diet is associated with a shift in the gut microbiota towards a more pathogenic/pro-inflammatory community.

Our cohort was primarily comprised of low-income Black and Latinx pregnant persons. Intake of highly processed foods is a hallmark of a Western diet, a diet pattern that is more common among disadvantaged minorities in the U.S., as these foods are more affordable and attainable for individuals with high financial burden (83). Previous studies from large perinatal cohorts, such as the 30-year longitudinal AVON study, have shown that women with lower access to high quality foods, have decreased vitamin and mineral intake (6). Our results support the hypothesis that poor diet quality is linked to insufficient vitamin and mineral dietary intake and accompanied by pro-inflammatory adjustments in the gut microbiome composition and metabolic structure.

### Strengths and Limitations

Our work focused on an understudied population at high risk of multiple health disorders, such as hypertension and GDM (6, 84). Associations between diet inflammatory potential and gut microbiota during pregnancy are under explored, and our research indicates that there is a significant link between microbial composition and metabolic functions and dietary inflammatory potential. Our work could be further improved by employing a more comprehensive dietary assessment approach that can assess all the 45 dietary parameters to calculate DII instead of just a portion of them (27 used for this study); including a larger sample size of a more diverse population in terms of DII scores that is followed longitudinally to determine the effects of DII on the gut microbiome later in pregnancy and perinatal disease development; employing a single stool sampling method; utilizing the same diet assessment for all participants and at the same collection time; employing sequencing technologies that enable to measure the abundance of microbial genes, such as shotgun sequencing (metagenomics), instead of relaying in metabolic predictions; and further characterizing the host immune and metabolic profiles.

## Conclusion

A proinflammatory diet, measured by DII, characterized by low intake of vitamins B12, B6, and A and iron, magnesium, niacin, riboflavin, and zinc, during early pregnancy is associated with a pro-inflammatory shift in the gut microbiota and metabolism as indicated by increase in galactose metabolism and methylglyoxal detoxification and multi drug efflux pump expression. Further characterization of gut metabolic status as a function of dietary alterations can provide opportunities for future research and targeted intervention strategies for at risk perinatal populations.

## Data Availability

The R notebook containing all analyses and de-identified data can be found at https://github.com/LabBea/Perinatal_DII
All data will be made available through the SRI repository.

https://github.com/LabBea/Perinatal_DII

## Abbreviations

DHQ-II: Diet History Questionnaire II
DII: Dietary Inflammatory Index
FFQ: Food Frequency Questionnaire

## Acknowledgements

LTH, SAA and BPB designed the current study; PM designed the original study; ESW, UN, BPB recruited participants and collected the samples; LBP and BL assessed diet using DHQII; SAA analyzed the data; SAA, LTH and BPB interpreted the data; SAA and BPB wrote the initial manuscript. All authors critically read and approved the manuscript. BPB was funded by the Arnold O. Beckman Postdoctoral Award, K12 BIRCWH Award (K12HD101373) and a NARSAD Young Investigator Award from the Brain and Behavior Research Foundation. JAG was funded by an NIDDK 1U24DK131617. SAA was supported by the University of Illinois Medical Scientist Training Program. This work has been also partially funded by the NICHD R03HD095056. REDCap application is supported though the Center for Clinical and Translational Science (CCTS) UL1TR002003.

## Reference Code

All analyses were performed in R. The R notebook containing all analyses and de-identified data can be found at https://github.com/LabBea/Perinatal_DII

## Data Availability

Data will be made available through the SRI repository.

## Sources of Support

R03HD095056

1U24DK131617-01 (JAG)

K12HD101373 (BPB)

Arnold O. Beckman Postdoctoral Fellowship (BPB)

NARSAD Young Investigator Award, Brain and Behavior Research Foundation (BPB)

University of Illinois Medical Scientist Training Program (SAA)

## Supplementary Materials

**Supplemental Table 1: DII differentially abundant ASVs using zero-inflated generalized linear models.** Corrected by subject age, gestational weeks, sample type, FFQ type, and BMI (adjusted p-value < 0.05).

*Attached excel file*

**Supplemental Table 2: DII differentially abundant microbial enzymes using zero-inflated generalized linear models**. Corrected by subject age, gestational weeks, sample type, FFQ type, and BMI. Microbial enzymes were all increased (N=38) by DII. We employed the KEGG database as a reference.

*Attached excel file*

**Supplemental Table 3: Microbial enzymes per term identified by Gene set enrichment by DII**. Microbially enzymes that were positively associated with DII before multiple comparison adjustments (N=194).

*Attached excel file*

**Supplemental Figure 1:**
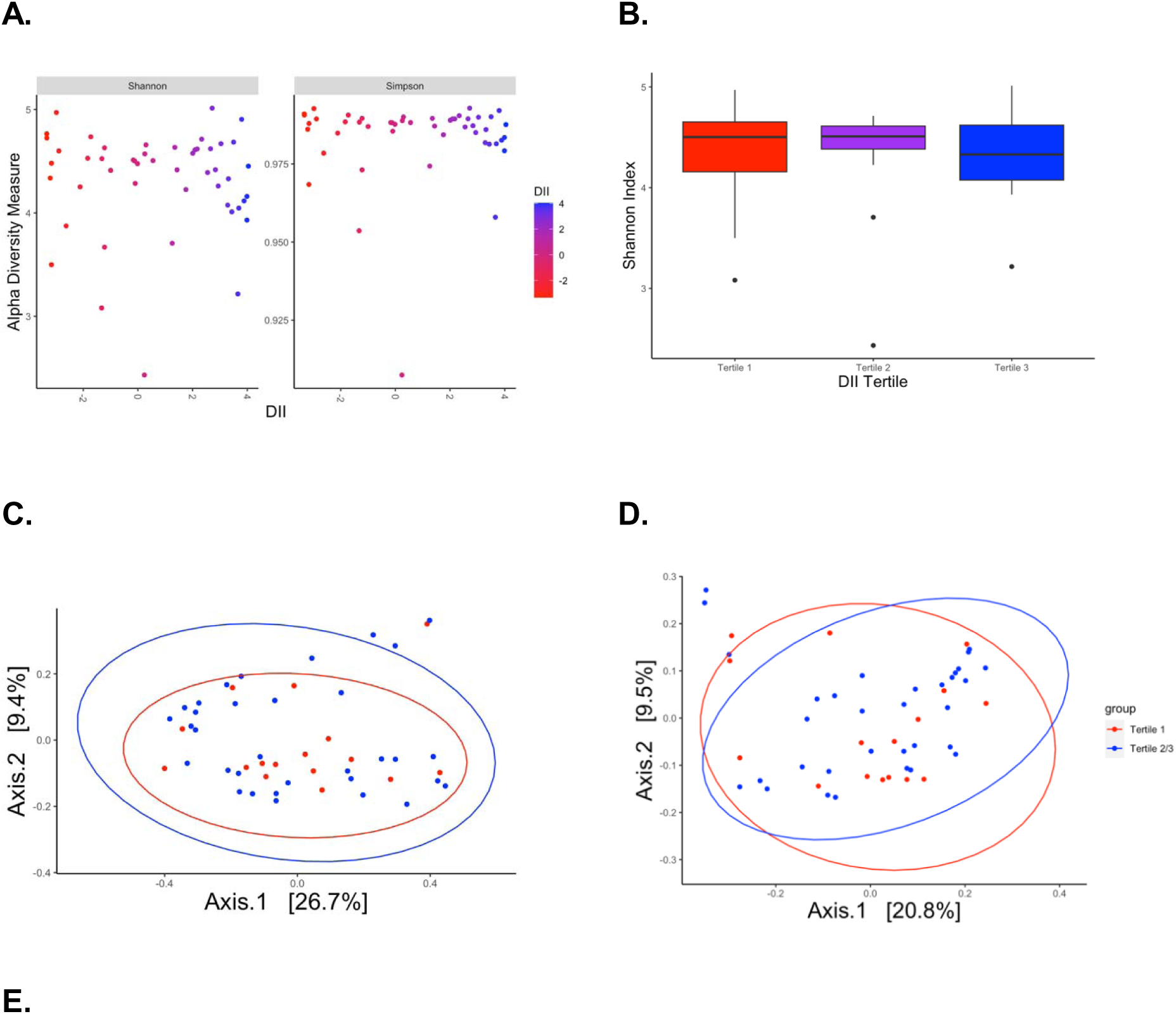
Alpha and beta diversity were not associated with assessment by DII score. **A:** Shannon and Simpson indexes as a function of DII scores. **B:** DII tertile (Wilcox Rank Sum p-value > 0.05). **C:** Beta diversity measured by Bray Curtis distance as a function of DII tertiles (PERMANOVA, p-value > 0.05). **D:** Beta diversity measured by UniFrac distance. Ellipses represent Tertile 1 and Tertile 2/3.

